# The Oral Fingerprint: Rapid comparison of palatal rugae for forensic identification

**DOI:** 10.1101/2025.05.30.25328631

**Authors:** Anika Kofod Petersen, Palle Villesen, Line Staun Larsen

## Abstract

The palatal rugae has been suggested to be just as unique as the human fingerprint. Therefore, endeavors have been made to utilize this uniqueness for identification of disaster victims. With the rise of digital 3D dental data, computational comparisons of palatal rugae have become possible. But a direct comparison of the full palatal scan by Iterative Closest Point (ICP) has shown to be tedious and demands knowledge of superimposition software. Here, we propose (1) an automatic extraction of the palatal rugae ridges from the 3D scans, followed by (2) ICP of the extracted ridges. Pairwise comparisons of palates takes less than a second, and in this study, it was possible to distinguish between palates from the same individual vs palates from different individuals with a ROC-AUC of 0.994. This shows that the extraction of the palatal rugae ridges is a potential efficient addition to the toolbox of a forensic odontologist for disaster victim identification.

## 1 Introduction

In the case of disasters, forensic odontology is applied in the identification of the disaster victims as one of the primary identifiers, alongside DNA analysis and fingerprints (1–4). In forensic odontology, information gathered from dental records is compared with the victim’s current dentition to get any insights into identification, i.e. any dental work, such as fillings or crowns, or any characteristic oral morphology (2–5). For these traits to aid in the identification of the individual, the sum of traits must be unique enough for the forensic odontologist to make a confident decision (3).

The palatal rugae, the ridges at the roof of the mouth, has been hypothesized to construct just as unique a pattern as a fingerprint, why this morphological structure is also considered for identification purposes (6–11). Even though this hypothesis has not been unanimously accepted, several studies point towards rugae uniqueness between individuals (6, 7, 9–11), even showing differences between monozygotic twins (12, 13).

This uniqueness has started endeavors to utilize the 3D landscape of palatal rugae for identification (6–8, 11, 13). Specifically, with the rise of 3D intraoral scanners (14–17), the palatal rugae can be scanned and represented as a 3D mesh. Several researchers have investigated different ways to compare 3D scans of palatal rugae, to aid in identification (6–8, 11–13, 18–23). Most of these studies cover superimposition methods (6, 8, 12, 13, 17, 18, 20–22), many being versions of the Iterative Closest Point (ICP) algorithm (6, 8, 12, 13, 17, 18, 20, 22), where the distances between one 3D mesh is minimized to the other to create a ‘best-fit’ overlay to then evaluate goodness of fit. When considering palatal rugae comparison, ICP has two major drawbacks (15, 24, 25). First, it is sensitive to the cropping of the palatal rugae (24, 25). If the tooth-gingiva border is meticulously traced when cutting out the rugae from an intraoral 3Dscan, there is a risk of ICP optimizing the fit of the dental traces, meaning the main driver for a good overlay may be tooth positions. This is not necessarily unwanted behavior, but in these cases, no guarantee can be made that the matching of the palatal rugae is what is being investigated. The other drawback is time, since ICP is an all-to-all comparison and a 3D scan typically harbors hundreds of thousands of datapoints (6, 13, 24, 25).

Even though superimposition techniques show good results, reaching an optimal overlay between two high resolution 3D surfaces is so time-consuming that it is difficult to apply in many scenarios (6, 13). For example, when considering a disaster victim identification case, where one 3D mesh of a palatal rugae surface is compared to a whole database of ante mortem 3D meshes. This process is then iterated for every victim of the disaster. Therefore, even though good results have been shown previously, there is a need for optimizing the superimposition method to make it feasible for palatal rugae comparison in forensic odontology disaster victim identification (6, 13).

One way is to not optimize the superimposition methods directly but instead preprocess the 3D scans to reduce the number of datapoints and only focus the process on the palatal rugae and not the rest of the palatal tissue. It has been suggested by Zhao et al. to manually extract the palatal rugae prior to ICP (20), but since this is both time consuming and add subjectivity in the data extraction step, an automated approach should be preferred. This study investigates the feasibility of automated extraction of the palatal rugae prior to ICP. This eliminates the problems associated with manual extraction of the palatal rugae while it retains the lower processing time and the unique traits of the palatal rugae (6, 13, 20).

## 2 Materials and methods

The palatal surface was manually cut (avoiding tracing the dentition border) from 102 intraoral scans of 51 healthy volunteering individuals, from a dataset previously presented by Kofod Petersen and colleagues (26). Each individual was subject to two intraoral scans approximately 6 months apart (26). Each scan was saved as a 3D surface mesh file in stl format (26).

To extract the palatal rugae from the intraoral scans, the scans were initially decimated using quadratic decimation to 50% resolution (27). Afterwards, the discrete mean curvature measurements were extracted from the surface mesh and the curvatures were normalized (27). The normalized curvature measures were then used to find the rugae ridges as follows. First, the 5% lowest curvatures, i.e. ‘the valleys’, were extracted to ensure that the border between the rugae ridge and the flat palatal soft tissue were included in the analysis. Followingly, the 30%, 25%, 20%, 15%, 10% and 5% highest curvatures, i.e. ‘the hills’, were extracted as point clouds. This means that each intraoral scan generated six files. This was done to investigate how much of the rugae ridges was needed for proper superimposition.

When reporting a similarity score, the score represents how well a pair of meshes align with one another. Having 51 individuals with 2 scans ensures that there are 51 matching pairs. To avoid biasing the reported performance, the dataset was randomly split into a validation set and a testing set. The validation set consisted of 26 of the matching pairs and 650 mismatching pairs (26*25). The testing dataset consisted of the remaining 25 matching pairs and 600 mismatching pairs (25*24).

Pairs of rugae 3D meshes were compared using ICP registration with 1000 maximum iterations and a maximum correspondence distance of 3 mm (28). This resulted in inlier root mean squared error (RMSE) measures for each comparison, describing the quality of alignment of the rugae meshes. A low inlier RMSE equals a good alignment. First, the inlier RMSE of all the matching pairs were compared to the inlier RMSE of the mismatching pairs of the validation dataset. To decide how much of the palatal rugae curvature to include different amounts of curvatures were tested. The amount of curvature that showed the highest Wasserstein distance between matching and mismatching pair inlier RMSE was chosen. If the method performs well, the matching pairs will have a low inlier RMSE, while the mismatching pairs will have a high inlier RMSE.

To report how well the method is at scoring the pairs appropriately, and to get an estimate of robustness of the procedure, we used a repeated sampling procedure (similar to bootstrapping) on the test set. For 1000 iterations, all matching pairs and 200 randomly selected mismatching pairs from the test dataset were compared. The relation between the true positive rate and the false positive rate was investigated using Receiver Operating Characteristic (ROC) area-under-the-curve (AUC).

## 3 Results

### 3.1 Defining the model using the validation set

The first step was to find an appropriate amount of rugae curvatures to include in a comparison. As seen in Figure 1, the largest distance between the matching and the mismatching inlier RMSE distributions was found when using the 5% lowest curvatures and the 10% highest curvatures.

**Figure 1.**
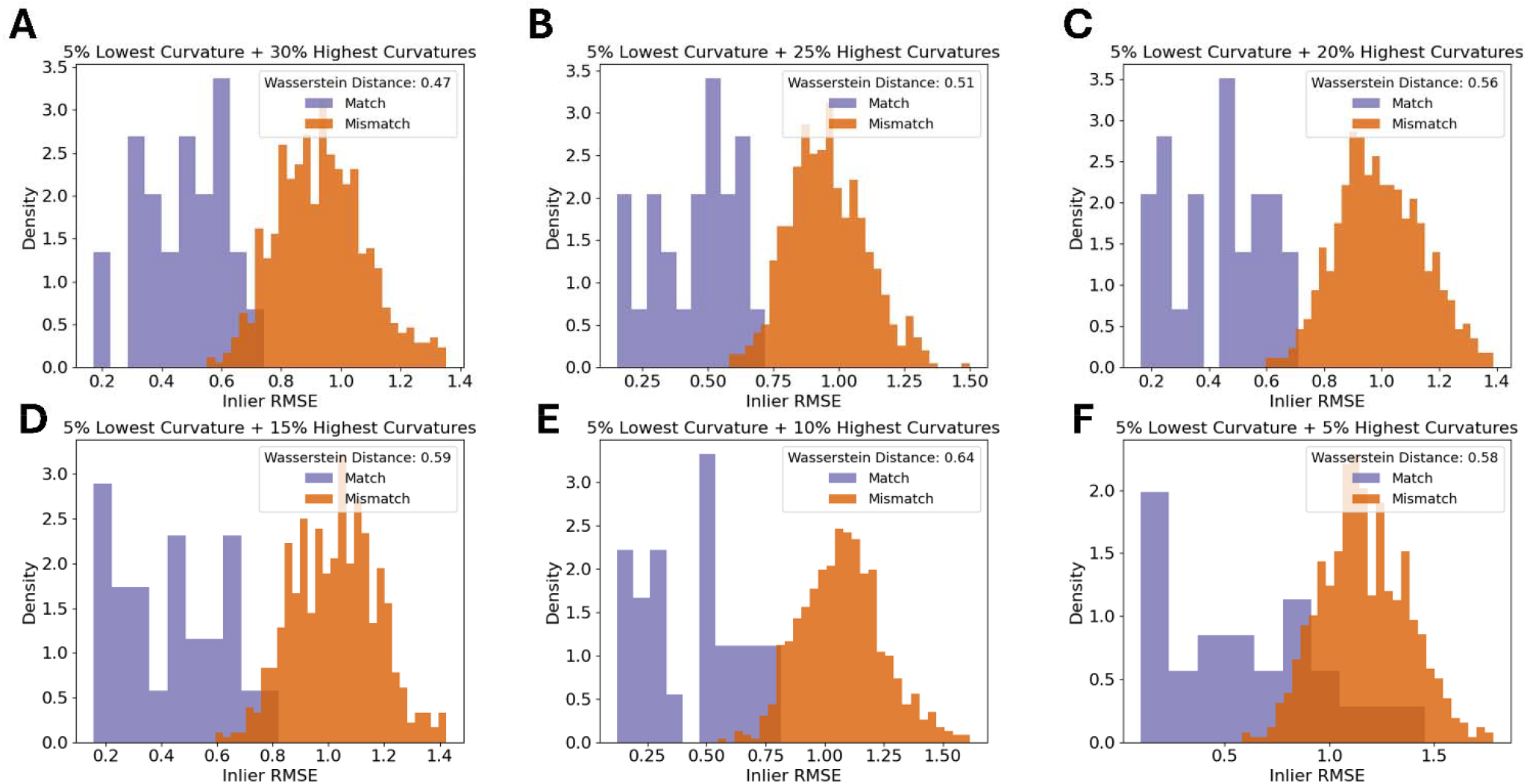
Density histograms of inlier RMSE of matching pairs and mismatching pairs for a decreasing amount of palatal curvature data. Figure shows the 5% lowest curvatures and the (A) 30% highest curvatures, (B) 25% highest curvatures, (C) 20% highest curvatures, (D) 15% highest curvatures, (E) 10% highest curvatures and (F) 5% highest curvatures,

When using 10% of the highest curvatures to calculate inlier RMSE, nearly all of the 26 rugae had the lowest inlier RMSE with the correct match (Table 1). In two cases the correct rugae was ranked as number 2. In these two cases, the difference in similarity score between the outranking mismatch and the match was 0.094 and 0.001 respectively, as seen in Table 1. In the cases where the match is the best ranking comparison, there was a mean difference between rank 1 and rank 2 of 0.393, indicating that a great difference between the best ranking and second-best ranking comparison is a good indicator of confidence.

**Table 1.** Lowest 6 Inlier RMSE scores for each query rugae in the validation dataset. True match indicated with bold and asterisk.

| **Rank 1** | **Rank 2** | **Rank 3** | **Rank 4** | **Rank 5** | **Rank 6** |
| --- | --- | --- | --- | --- | --- |
| **0.305\*** | 0.638 | 0.878 | 0.918 | 0.924 | 0.931 |
| **0.216\*** | 0.733 | 0.779 | 0.789 | 0.837 | 0.877 |
| **0.606\*** | 0.949 | 0.964 | 1.028 | 1.035 | 1.053 |
| **0.687\*** | 0.872 | 1.038 | 1.125 | 1.127 | 1.137 |
| 0.685 | **0.778\*** | 0.836 | 0.98 | 0.991 | 0.992 |
| **0.529\*** | 0.751 | 0.791 | 0.819 | 0.833 | 0.842 |
| **0.640\*** | 1.038 | 1.106 | 1.106 | 1.111 | 1.118 |
| **0.659\*** | 0.833 | 1.067 | 1.069 | 1.094 | 1.094 |
| **0.394\*** | 0.636 | 0.790 | 0.798 | 0.802 | 0.813 |
| **0.816\*** | 0.919 | 1.054 | 1.134 | 1.169 | 1.197 |
| **0.122\*** | 0.649 | 0.807 | 0.816 | 0.819 | 0.820 |
| **0.296\*** | 0.796 | 0.834 | 0.855 | 0.871 | 0.898 |
| **0.265\*** | 0.725 | 0.741 | 0.76 | 0.763 | 0.798 |
| **0.142\*** | 0.739 | 0.803 | 0.809 | 0.826 | 0.826 |
| **0.221\*** | 0.803 | 0.833 | 0.843 | 0.898 | 0.911 |
| 0.687 | **0.689\*** | 0.707 | 0.775 | 0.779 | 0.832 |
| **0.188\*** | 0.546 | 0.745 | 0.8 | 0.804 | 0.856 |
| **0.486\*** | 0.938 | 0.961 | 0.998 | 1.008 | 1.067 |
| **0.571\*** | 0.845 | 0.896 | 0.986 | 1.020 | 1.070 |
| **0.508\*** | 0.825 | 0.877 | 0.895 | 0.948 | 0.975 |
| **0.478\*** | 0.802 | 0.881 | 0.884 | 0.918 | 0.921 |
| **0.224\*** | 0.964 | 1.034 | 1.041 | 1.052 | 1.056 |
| **0.497\*** | 0.875 | 0.877 | 0.916 | 0.921 | 0.942 |
| **0.294\*** | 0.763 | 0.785 | 0.862 | 0.877 | 0.897 |
| **0.536\*** | 0.891 | 0.922 | 0.958 | 0.986 | 1.005 |
| **0.169\*** | 0.758 | 0.829 | 0.829 | 0.838 | 0.858 |

### 3.2 Testing the scoring

The inlier RMSE scoring was tested on the test data with palatal rugae from the 10% highest curvatures (and the 5% lowest curvatures) on the remaining 25 palatal meshes. Following the same procedure as for the validation dataset, we saw that 24/25 of the matches were found to have the lowest inlier RMSE (Table 2). The single false negative had the correct match ranked 6, but with an overall high RMSE.

**Table 2.** Lowest 6 Inlier RMSE scores for each query rugae in the test dataset. True match indicated with bold and asterisk.

| **Rank 1** | **Rank 2** | **Rank 3** | **Rank 4** | **Rank 5** | **Rank 6** |
| --- | --- | --- | --- | --- | --- |
| **0.536\*** | 0.707 | 0.725 | 0.768 | 0.826 | 0.829 |
| **0.849\*** | 0.927 | 0.952 | 0.955 | 0.966 | 0.978 |
| **0.681\*** | 0.823 | 0.852 | 0.882 | 0.889 | 0.954 |
| 1.091 | 1.112 | 1.209 | 1.234 | 1.285 | **1.291\*** |
| **0.273\*** | 0.84 | 0.843 | 0.854 | 0.961 | 0.984 |
| **0.253\*** | 0.662 | 0.685 | 0.787 | 0.792 | 0.857 |
| **0.852\*** | 1.083 | 1.083 | 1.102 | 1.146 | 1.158 |
| **0.356\*** | 0.733 | 0.887 | 0.894 | 0.907 | 0.911 |
| **0.294\*** | 0.983 | 1.015 | 1.049 | 1.06 | 1.063 |
| **0.534\*** | 0.855 | 0.861 | 0.864 | 0.868 | 0.878 |
| **0.595\*** | 0.845 | 0.966 | 0.982 | 0.992 | 0.999 |
| **0.674\*** | 0.91 | 0.977 | 0.978 | 1.031 | 1.068 |
| **0.761\*** | 0.787 | 0.822 | 0.837 | 0.861 | 0.903 |
| **0.499\*** | 0.757 | 0.772 | 0.793 | 0.811 | 0.818 |
| **0.548\*** | 0.725 | 0.75 | 0.805 | 0.827 | 0.868 |
| **0.155\*** | 0.784 | 0.794 | 0.85 | 0.854 | 0.854 |
| **0.449\*** | 0.828 | 0.836 | 0.889 | 0.919 | 0.959 |
| **0.403\*** | 0.86 | 0.953 | 1.012 | 1.023 | 1.053 |
| **0.495\*** | 0.802 | 0.861 | 0.92 | 0.939 | 0.94 |
| **0.613\*** | 0.726 | 0.75 | 0.781 | 0.817 | 0.825 |
| **0.712\*** | 1.031 | 1.053 | 1.061 | 1.066 | 1.109 |
| **0.853\*** | 0.882 | 0.976 | 1.002 | 1.012 | 1.152 |
| **0.415\*** | 0.648 | 0.675 | 0.731 | 0.759 | 0.776 |
| **0.127\*** | 0.784 | 0.804 | 0.832 | 0.853 | 0.872 |
| **0.324\*** | 0.723 | 0.816 | 0.818 | 0.87 | 0.891 |

We found the scoring procedure to have an extremely high performance with a mean ROC-AUC of 0.994 (Figure 2). The 1000 random subsamples also showed the procedure to be extremely robust with AUCs ranging from 0.988 to 0.998 (Figure 2, shaded band).

**Figure 2.**
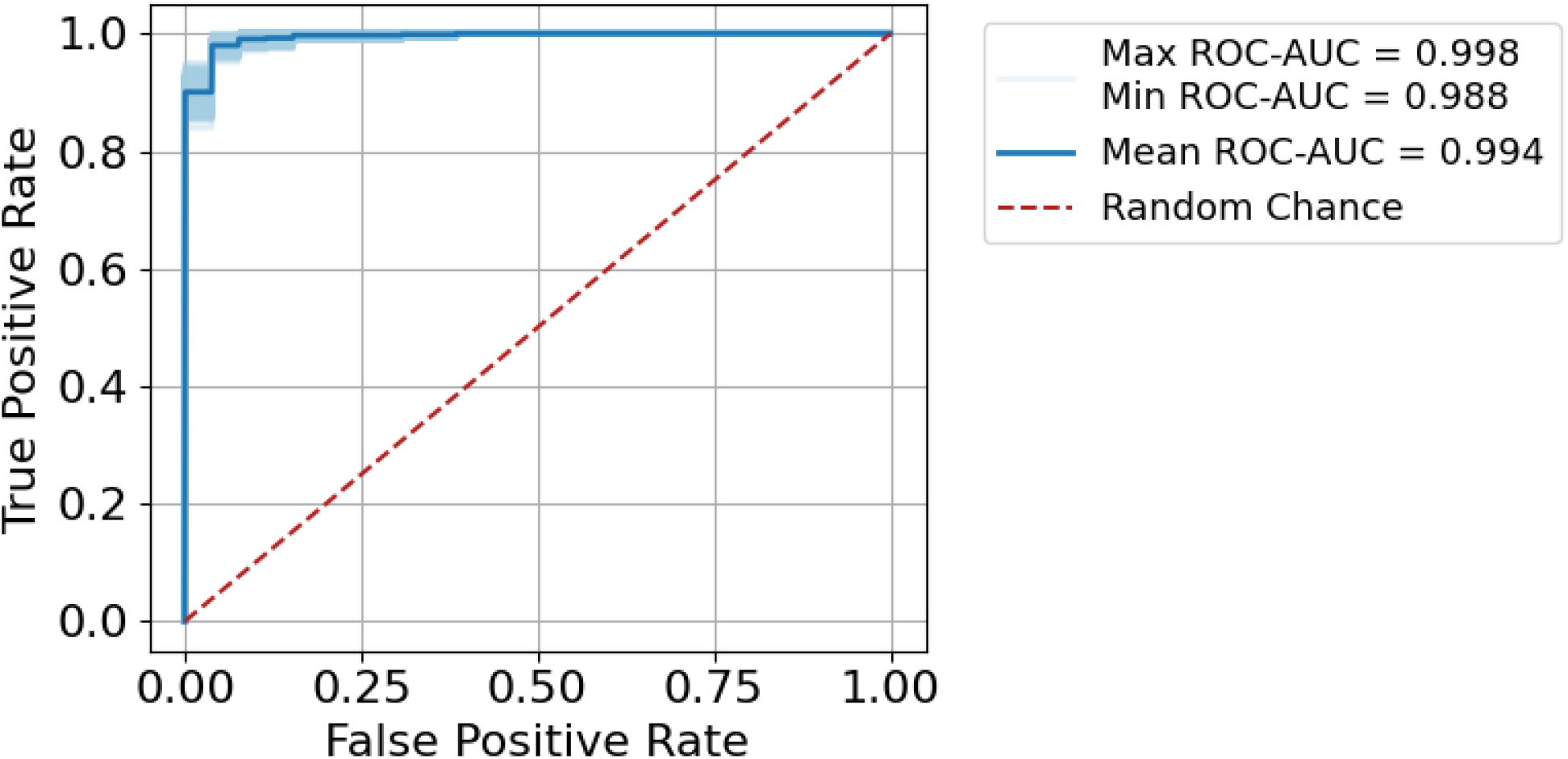
ROC-AUC for the test data using 1000 iterations of subsampling.

### 3.3 Processing time

Extracting the palatal curvatures and saving the rugae files are only done once per intraoral scan and can easily be done in parallel. Therefore, this part of comparison is not expected to be time limiting. In contrast, since ICP is done for all victims across all *ante mortem* 3D meshes, the number of ICP comparisons will significantly increase with the number of victims and the size of the *ante mortem* database.

On a machine with an 11th Gen Intel Core i7-1165G7, 2.80GHz (x86_64) processor and 8GB RAM, a comparison of one pair of extracted rugae curvatures took on average 0.078 seconds (PI 95% from 0.011 seconds to 0.251 seconds). To perform the same analysis on full palatal rugae scans took on average 47.9 seconds (PI 95% from 8.24 seconds to 157.27 seconds).

For exemplification, 500 comparisons were made where 51 were matches and the remaining 449 were mismatches. To perform ICP on only the 500 extracted rugae comparisons took a total of 43 seconds, while ICP on the 500 full palatal comparisons took a total of 6 hours 39 minutes and 34 seconds. Despite the higher amount of data, the performance was worse when using the full palatal meshes (Figure 3). It seems that the extra surface information and the difference in mesh border might drive the ICP alignment away from the optimal rugae alignment, making automatic ICP registration of full palatal meshes difficult, resulting in poor performance (15, 25, 26).

**Figure 3.**
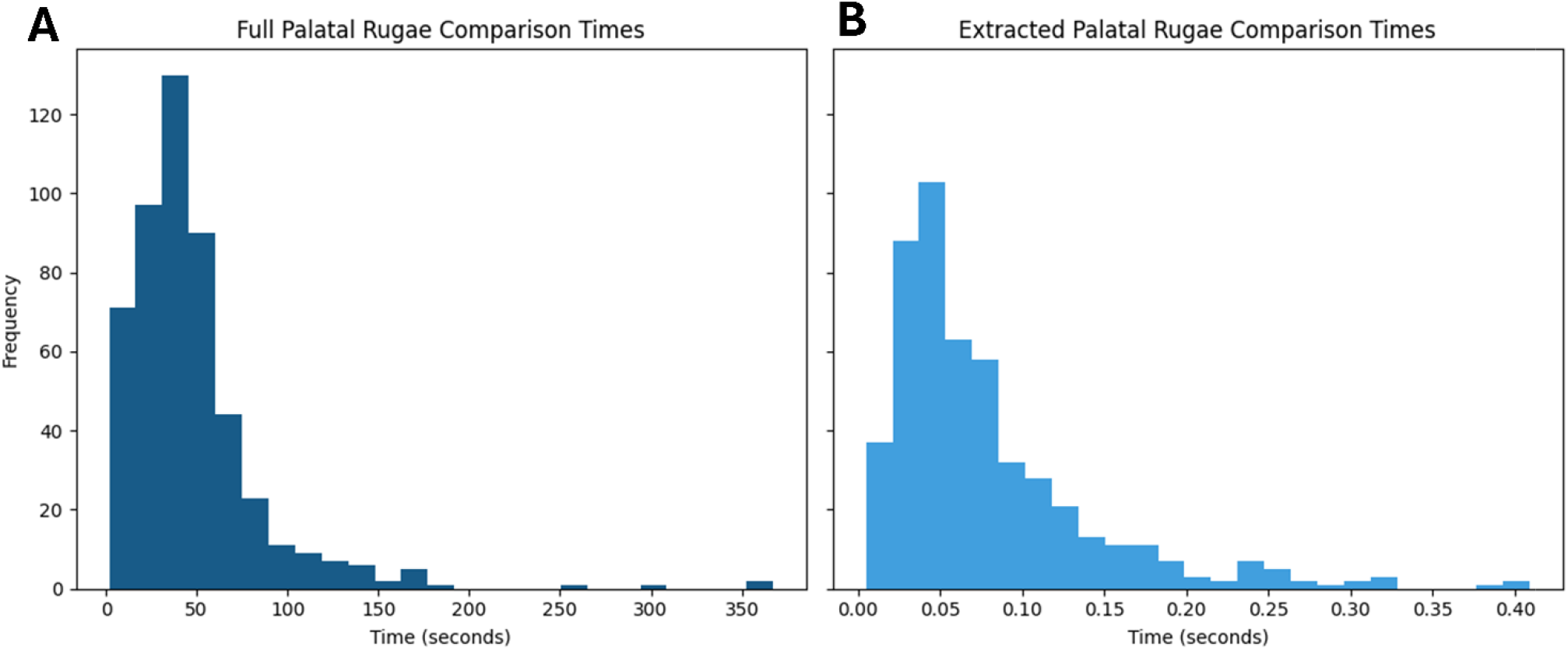
Histograms of comparison times for (A) full palatal rugae and (B) extracted palatal rugae.

## 4 Discussion

For disaster victim identification using Forensic Odontology, an automated workflow for palatal rugae comparison in 3D could be beneficial (6–8, 13, 14, 19–23). But with the preferred methodology (ICP) being too slow when it comes to comparing full palatal meshes, this is currently not feasible for major disasters (6, 13).

However, by automatically extracting the 10% highest curvatures and the 5% lowest curvatures from the palatal rugae scans, a comparison can be done in less than a second, which is no longer a bottle neck when it comes to disaster victim identification. This data preprocessing step increases the speed of comparisons by magnitudes, alleviating the time constraint.

Furthermore, the extraction of rugae curvatures ensures that the optimal alignment is focused on the rugae ridges and no other features of the palatal surface, such as cropping border and tooth position.

The higher level of detail in the full palatal meshes does not increase separability between matches and mismatches. Rather the contrary, as seen when comparing Figure 1 and Figure 4. We see an increase in speed and separation when using only the extracted palatal curvatures, highly advocating for the use of extracted palatal rugae instead of full palatal meshes.

**Figure 4.**
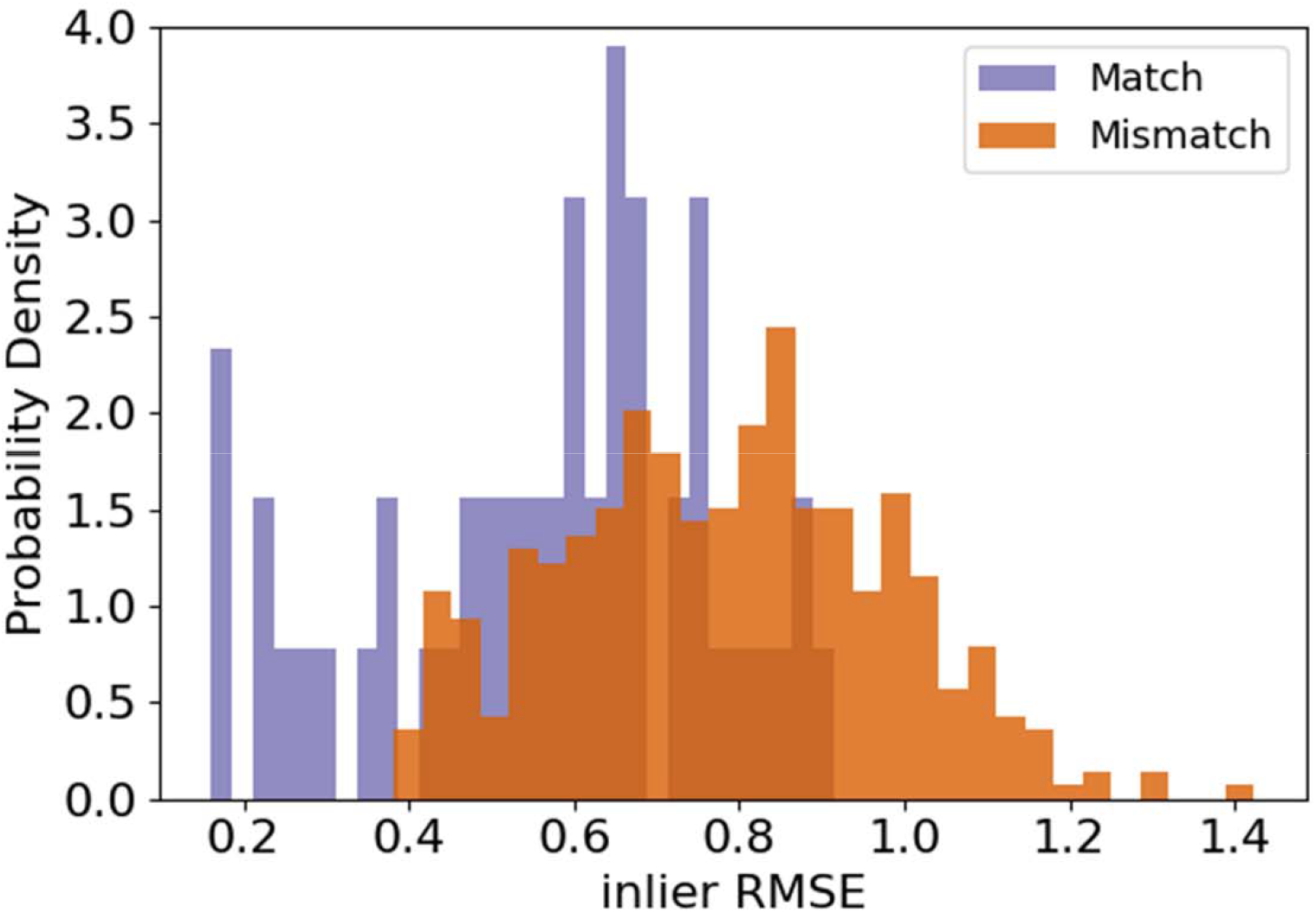
inlier RMSE after ICP on full palatal rugae for matches and mismatches.

To utilise rugae uniqueness for identification purposes, there is a need for preserved rugae patterns. This means that in DVI scenarios where the palatal soft tissue has suffered great damage, e.g. by heat or decomposition, this rugae comparison methodology would not be feasible. But if the palatal rugae are intact, they can serve as an addition to other identification methods, like dental comparison, DNA and fingerprints, now without being a time limiting step.

This study shows that automatic extraction of palatal rugae from 3D intraoral scans can be used in combination with ICP for disaster victim identification, without causing a major bottleneck due to processing time (6–8, 12, 13, 18–23). The inlier RMSE of the extracted palatal rugae can serve as a similarity score, used for ranking possible matches. The ranking of the palatal rugae comparison can then be used in addition to other forensic odontological techniques for the forensic odontologist to identify disaster victims (3).

## Author Contributions

AKP: Conceptualization, Methodology, Formal analysis, Investigation, Data Curation, visualization, Writing - Original draft, Writing – Review and editing, PV: Conceptualization, Writing – Review and Editing, Supervision LSL: Conceptualization, Writing – Review and Editing, Supervision, Funding acquisition

## Acknowledgments

We thank Professor Emerita, Dorthe Arenholt Bindslev, for her valuable discussions and insightful input during the conceptualization phase of this work.

## Data Availability Statement

This study includes data that can be considered personal dental data, which the authors are not authorized to share.

## Ethics Statement

This study was registered with the Data Protection Unit at Aarhus University, Denmark (file number 2016-051-000001, serial number 2534; file number 20220367531, serial number 3155). It was conducted in full accordance with the World Medical Association Declaration of Helsinki and complied with the European Union General Data Protection Regulation legislation. According to the assessment by the chairmanship of The Danish National Committee on Health Research Ethics (NVK), the study and its protocol are deemed exempt from notification (case number: 2400741) thus approving the protocol.

